# RELATIONSHIP BETWEEN ADVERSE EVENTS PREVALENCE, PATIENT SAFETY CULTURE AND PATIENT SAFETY PERCEPTION IN A SINGLE SAMPLE OF PATIENTS: A CROSS-SECTIONAL AND CORRELATIONAL STUDY

**DOI:** 10.1101/2021.03.25.21254370

**Authors:** Mónica Chirinos, Carola Orrego, Cesar Montoya, Rosa Suñol

## Abstract

**Objective:** To assess the relationship between Adverse Events Prevalence, Patient Safety Culture and Patient Safety Perception.

**Design:** Cross-sectional, ex post facto comparative study on a single sample of patients.

**Setting:** Four medium-high-level hospitals were included in the study — two public and two private from Zulia State in Venezuela.

**Participants:** 556 medical records and patients were studied for the prevalence and patient safety perception study, and 397 of the healthcare providers involved in the care of these patients were surveyed for the patient safety culture study, at two public and two private hospitals.

**Outcome measurement:** The primary outcome of this study was the Association between Adverse Events Prevalence, Patient Safety Culture and Patient Safety Perception, and according to hospital funding type, private and public.

**Results:** An inverse association was observed between Adverse Events Prevalence and its severity and Patient Safety Culture Index (PSCI: rho=-0.8 p=0.5) (95% CI=0.26-0.10) and Patient Safety Perception Index (PSPI: rho=-0.6 p=0.18) (95% CI=0.10-0.28), which were protective factors for patient safety. No association was identified between Patient Safety Culture and Patient Safety Perception (rho=0.0001). No statistical differences were identified by hospital type (p=0.93) (95% CI=0.70-1.2).

**Conclusions:** The analysis of the variable correlations studied (AEP, PSC and PSP) within the same sample offers an interesting and useful perspective. In this sample, although no correlation was observed between the three variables as an interacting set, some correlation patterns were observed between pairs of variables that could guide further studies.

**Strengths and limitations of this study:**

- The association analysis of the three main variables (adverse events prevalence, patient safety culture and patient safety perception) in the same sample of patients and health professionals and over the same time period during which the care process and, consequently, the adverse events took place is an unprecedented and innovative approach, which contributes to the body of knowledge on this field of study.
- The relational theoretical framework produced provides a “map” that can facilitate decision-taking and which establishes the variables that should be considered a priority. As a result, it is highly likely to have an impact on the clinical efficiency and safety of healthcare institutions and services.
- The study was carried out at both private and public hospitals, enabling the behaviour of the aforementioned variables to be compared and analysed at each type of institution. The literature to date on this approach is scarce and offers little information.
- Although the sample of hospitals selected is not representative of Venezuela, it is representative of the hospitals of one of the most productive and populated states in the country.
- During stages 1 and 2, some resistance to collaborating with and completing the evaluation instruments was identified among the health professionals.

## INTRODUCTION

Patient safety has been widely studied from the perspective of adverse events (AEs) and their financial impact, the quality/safety culture at institutional level and patients’ perceptions of AEs and how they are managed. Other studies in this field have adopted varied approaches in terms of their definitions of AEs, purpose,(1,2) objectives and design elements,(3,4) whether based on medical records (retrospective studies and cross-sectional studies),(5–7) or on individual follow-up care (prospective studies).(8)

The literature in this field illustrates the broad research that has been carried out offering different perspectives on patient safety and consolidating the recommendation that patient safety culture (PSC) must be fostered by healthcare teams to minimize the risks inherent in clinical practice and prevent potential AEs.(9) In addition, the World Health Organization (WHO) has emphasized the importance of patient safety perception (PSP) by patients, in terms of learning from the unique perspective of those experiencing the complex emotions caused by a health condition.(10) Its role has been considered crucial for the sustainability of quality systems and patient safety in healthcare institutions and in society as a whole.(11) Furthermore, the Latin American Study of Adverse Events (IBEAS), published in 2008 to promote patient safety in the region, assessed AE incidence in hospitals in five Latin American countries and concluded that there was a global prevalence for patients with some kind of AE (10.5%) (CI=95% (9.91 to 11.04)).(7). According to the authors of the IBEAS, higher AEP can be linked to the complexity of healthcare systems in developing economies and, specifically, to a lack of material or poor healthcare infrastructure.(7,8) These findings support the idea that this type of research needs to be conducted on a wider scale in order to identify determining factors and develop appropriate policies to deal with AEs.

Although the literature shows studies that analyse the culture of patient safety, the prevalence and incidence of AEs, and the perception of safety by patients(12,13), all in isolation and individually or in a combination of two of the variables, we were unable to identify any studies that jointly evaluate these three variables within the same sample and over the same period of time.

Studies therefore need to be carried out at hospitals in different countries to analyse these interactions using a single sample of patients and healthcare providers directly involved in the care of those patients.

Another factor analysed in this study was patient safety according to hospital funding type; this aspect was considered important, since in many Latin American countries, including Venezuela, the health system is made up of a public sector, funded by the State, and a private sector funded by companies or directly by users.(14) Some authors argue that as a result of market competition within the private sector itself, it makes a greater effort than the public sector to guarantee safe and quality processes and is more attentive to the needs of patients.(15) An additional incentive in this respect is the higher risk of medical liability lawsuits. Research indicates that there is often a significant differentiation between public and private procedures in terms of patient safety and clinical responsibility,(16) so we also intend to make a contribution to this area of study.

The objective of this research is to describe the correlation model of AEP, PSC and PSP for the same population of hospitalized patients and their providers using existing validated instruments.

The separate results of these evaluations have been reported in detail in other papers;(17)(18) however, some of the descriptive elements have also been included here to aid comprehension.

## METHODS

This is a cross-sectional, comparative, and observational study, based on the simultaneous analysis of three variables: AEP,(17) PSC,(18) and PSP in a single sample of four hospitals during 2017, each of which are studied in three different investigations. 4 medium-high-level hospitals were included in the study — two public and two private(14). The analysis was conducted through a systematic and structured review of medical records and questionnaires that were distributed to health professionals and patients.

The medium-high complexity hospitals of the state of Zulia (this type of hospital offers four basic medical specialties plus four subspecialties, some specialized care units, and some treatment units that serve as a regional or national reference) were considered as the population, which represent a set of 13 hospitals in the region. Due to the geographical dispersion of the hospitals, multistage cluster sampling with stratification was used, proceeding as follows:

I. The hospitals sampled were taken from randomly selected, medium-high complexity hospitals on the east coast, represented by two hospitals (50% of the total), and on the west coast, also represented by two hospitals (31% of the total).
II. Secondary Sampling Units: Within the four established hospitals, the total number of patients to be studied was then determined by applying the proportion formula for a finite number of individuals (N=118), calculated in accordance with the total number of monthly hospital admissions (N=3000). The patients had to meet certain criteria for inclusion in the study: *cases with a minimum of 72 hours since admission into in-patient wards, *patients admitted into in-patient wards of the healthcare units of Internal Medicine, General Surgery, Traumatology and Orthopaedics, Gastroenterology, Nephro-Urology and Paediatrics. At each hospital, the patients were selected sequentially at random over a 30-day period, until the minimum estimated number of patients was reached, resulting in a sample of 139 per hospital and a total of 556 patients between the four hospitals.

The instruments were applied in order by following a logical and chronological sequence for obtaining the data, that is, first, the screening for AEs, second, patients’ perception, and third, the health professionals’ perception of PSC. The global study was carried out in three stages. During the first stage the patients were selected and the medical records were reviewed for the analysis of AEP. In this phase, both the patient sample and the health professionals who intervened in the processes of these hospitalized patients, as described above, were determined. In the second stage of the investigation, the patients still hospitalized were analysed to determine their perception of their clinical safety. In the third stage, the culture of patient safety was studied by surveying the professionals who intervened in the process of caring for those same patients. The methodology used in each of the studies that form part of this research is briefly described below:

I.- Determining AEP and AE severity:(17) The aim of this particular study was to determine and describe AEP in hospitals in the state of Zulia. A total sample of 556 hospitalized patients was selected randomly over four months. The same methodology and definition established in the Harvard study on AE incidence, which are published in detail in another paper, were implemented and applied in two phases:(1) 1. Screening of AEs: this was conducted by applying the screening guide for AEs and the clinical history form, both of which had been adapted and validated.(1,5,7,19). In the few cases where the patient was visited by the health professional, this took place due to insufficient information in the clinical history to detect and/or characterize the AE. This occurred at the end of the patient’s hospital stay or after completion of the PSP questionnaire, to avoid any bias.

2. Detection of AEs: this was conducted using the Spanish version of the MRF2 questionnaire (Modular Revised Form) for case reviews.(1,5,16,19).

### Variables analysis

Percentage of AEs by hospital refers to the total number of patients who experienced an AE; percentage of AEs by severity refers to the classification of severity levels into mild, medium and severe; and percentage of preventable AEs refers to the assessment by the evaluator of the clinical history of whether there was any evidence that the AE could have been avoided, using a qualitative 4-level scale divided into (i) slight possibility of prevention, (ii) moderate possibility, (iii) high possibility, and (iv) full evidence of prevention.

The Mann-Whitney U test was used on independent samples in order to ensure comparability between public and private hospitals, and the main result was that 93 patients had experienced some kind of AE, with an overall rate of 16.7% (CI = 95%), 20% in public hospitals and 18% in private hospitals. 80% of these AEs were determined to be avoidable, 30.1% were of low severity, 40.8% medium and 29% high.

### II. Determining the safety culture perceived by health professionals(18)

The aim of this study was to analyse predictors of PSP in public and private hospitals and examine the factors that contribute to it, constructing a new and specific theoretical and methodological model.

A survey was conducted among all the health professionals (N = 588) at the four hospitals involved in the care process of each patient in the AEP study. These professionals were identified by examining the patients’ medical records (medical specialists, nurses, nursing assistants, nursing technicians, hemotherapy specialists, nutritionists, laboratory assistants, bioanalysts, radiology technicians, social workers, graduate medical residents, administrative staff, and managers). The survey was carried out using a questionnaire entitled “Analysis of patient safety culture in hospital environments”, based on the questionnaire used in the Agency for Healthcare Research and Quality (AHRQ) Hospital Survey on Patient Safety Culture,(20,21) and adapted to the Spanish context.(22)

A Patient Safety Culture Index (PSCI) was created and categorized into four levels to compare the PSC with the other previously established variables: Favourable (104-85), Moderately Favourable (84-65), Moderately unfavourable (64-45), Unfavourable (44-25).

The PSCI was calculated by applying factor analysis to the set of 12 dimensions covered in the instrument, as well as to the socio-professional characteristics and the additional information (score awarded to the degree of patient safety in the service/unit and the number of AEs reported, which was obtained through a Kaiser-Mayer-Olkin (KMO) value above 0.75 and a high level of significance in Bartlett’s test of sphericity).

To obtain a simple expression of the culture variable, all 12 dimensions of the instrument were regrouped, taking into account their related nature, into three factors, which were later combined to form the index.(22) The PSCI was calculated based on the following factors: occupational, communication, and organizational.(23) The analysis showed that all hospitals had a “moderately unfavourable” PSCI.

### III. Determining the PSP by patients

The objective of this study is to determine the level of perception of patients of their safety during the hospitalization process.

The PSP was calculated using a survey conducted by specifically trained data collectors (social workers) the day before the discharge of each of the 556 patients evaluated for the AEP study (139 patients at each hospital, with a 100% response rate). The survey was carried out by applying the previously validated instrument entitled “Questionnaire on perceived healthcare safety in the hospital environment” (QPHS).(24).

### Variables analysis

We used Perceived Adverse Events, which refers to those AEs, injuries, damages, and situations that the patients perceived to be a negative consequence of unsafe care or a complication other than that caused by their disease.

To obtain the correlation between the study variables, a Patient Safety Perception Index (PSPI) was created using the dimensions that make up this variable, as expressed in the measuring instrument (items established). To obtain a single statistical value and determine the index, the items were given a weighting with a score; these scores were subsequently added up. A PSCI was created and categorized into four levels to compare PSP with the other previously established variables: High (38-33), Moderately High (33-28), Medium-low (28-23), Low (23-18).

To compare the level of safety perception between the public and private hospitals, the Student t-test was used on independent samples. The main result is that a low index was obtained on the patients’ perception of their safety (PSPI PrH = 27.3 PuH = 25.4).

### IV. Evaluation of the association of variables

Spearman’s Rho was used as a statistical model to determine the strength and nature of the relationship between the PSCI and PSPI based on AEP and AE severity, regardless of the type of hospital.

### Patient and public involvement

During the conducting of this research, two variables were considered to directly involve patient participation:

1. AEP: During the AE screening process, on occasion the evaluating health professional had to delve more deeply into the case and visit the patient to discuss the purpose of the study and the visit.
2. Patient perception of safety: This variable was evaluated primarily by applying a previously developed and validated instrument, and therefore the patient did not participate in its design. The interviewer approached the patient, or occasionally a family member, in a timely and polite manner.
3. PSC: the opinions of health professionals involved in patient care were studied by providing them with the previously validated and prepared instrument for 48 hours; they were not involved in designing the questionnaire.

The results of the investigations that precede the present one have been presented and discussed at institutional and academic events to which the patient organizations of the hospitals participating in this study were invited.

## RESULTS

### I. Characteristics of the variables

The characteristics of the individuals observed are shown in Tables 1 and 2.

**Table 1.** Characteristics of the patient population presenting AEs, by hospital type.

| CHARACTERISTICS | PUBLIC HOSPITALS |  | PRIVATE HOSPITALS |  | TOTAL |  |
| --- | --- | --- | --- | --- | --- | --- |
|  | (PuH). |  | (PrH) |  |  |  |
| Gender | No. | % | No. | % | No. | % |
| Female | 41 | 44.0 | 30 | 32.2 | 71 | 76.3 |
| Male | 12 | 12.9 | 10 | 11.8 | 22 | 25.8 |
| Total | 53 | 57.0 | 40 | 43.0 | 93 | 100 |
| Age (years) | No. | % | No. | % | No. | % |
| less than 15 years | 1 | 1.0 | 0 | 0 | 1 | 1,2 |
| 16-25 years | 11 | 11.8 | 4 | 4.3 | 15 | 15.2 |
| 26-45 years | 21 | 22.5 | 18 | 19.3 | 39 | 42.4 |
| 46-65 years | 14 | 15.0 | 14 | 15.0 | 28 | 30.4 |
| > 66 years | 6 | 6.4 | 4 | 4.3 | 10 | 9.9 |
| Total | 53 | 57.0 | 40 | 43.0 | 93 | 100 |
| Attended by speciality | No. | % | No. | % | No. | % |
| Medicine (Non-surgical) | 11 | 11.8 | 8 | 8.6 | 19 | 20.4 |
| Paediatrics | 13 | 13.9 | 1 | 1.0 | 14 | 15.0 |
| Surgery | 12 | 12.9 | 8 | 8.6 | 20 | 21.5 |
| Traumatology | 3 | 3.2 | 2 | 2.1 | 5 | 5.3 |
| Nephrology | 3 | 3.2 | 4 | 4.3 | 7 | 7.5 |
| Other units | 11 | 11.8 | 17 | 18.1 | 28 | 30.1 |
| Total | 53 | 57.0 | 40 | 43.0 | 93 | 100 |
| Education level | No. | % | No. | % | No. | % |
| Illiterate | 6 | 6.4 | 0 | 0 | 6 | 6.4 |
| Primary | 18 | 19.3 | 7 | 7.5 | 25 | 26.8 |
| Secondary | 20 | 21.5 | 17 | 18.2 | 37 | 39.7 |
| Technical University | 4 | 4.3 | 5 | 5.3 | 9 | 9.6 |
| High University | 5 | 5.3 | 11 | 11.8 | 16 | 17.2 |
| Total | 53 | 100 | 40 | 100 | 93 | 100 |

Source: Chirinos, M. et al.

**Table 2.**
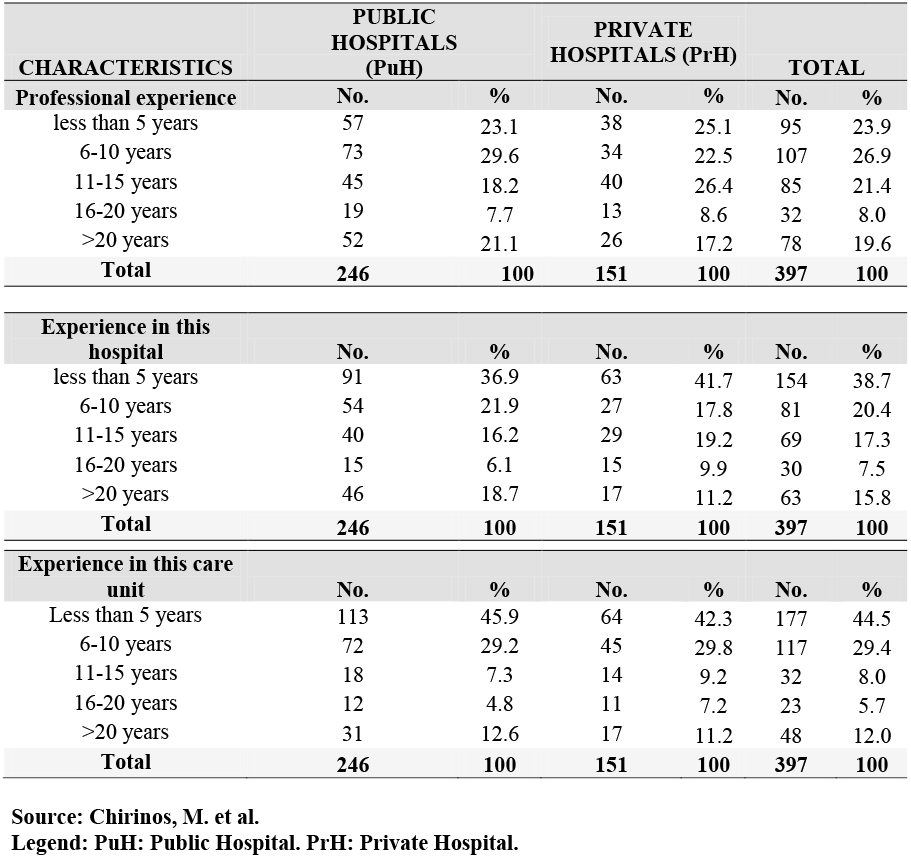
Characteristics of the healthcare provider population studied, by hospital type.

For both types of hospitals (Public Hospitals = PuH and Private Hospitals = Pr), the female population is higher than the male population, as is the 26 to 45 year-old age group with primary and secondary education levels for public hospitals, and with secondary and university education levels for private hospitals. Among the different care units, internal medicine and paediatrics account for the largest population in the private hospitals, with the addition of general surgery in the case of the public hospitals (see Table 1).

The response rate of the survey among health professionals was 68.8% (405 of the 566 professionals who were sent the survey responded), of whom 51% (N = 206) worked in public hospitals and 49% (N = 199) in private hospitals.

As shown in Table 2, which presents the most relevant characteristics of the population of health professionals who participated in the study, those with between 6 and 10 years’ professional experience are most highly represented; however, those with less experience, that is, less than 5 years, are also highly represented. Similarly, the health professionals most highly represented were those with less than 5 years’ experience both in the hospital and in the unit where they were currently working.

### II. Comparison of AEP, AE severity, PSC and PSP between the public and private hospitals

The prevalence and severity of AEs in both types of hospital did not show any significant statistical differences (Sig = 0.11 and a value of rho=0.66); the prevalence of moderately severe AEs is equally present in both types of institution, regardless of the type of hospital management (see Table 3).

**Table 3.** Hypothesis testing for the comparison of the three variables studied, by hospital.

| Table 3 |  |  |  |  |
| --- | --- | --- | --- | --- |
| VARIABLE | PUBLIC HOSPITALS | PRIVATE HOSPITALS | TOTAL | SIGNIFICANCE TEST |

| AEP | No. (%) | No. (%) | No. (%) | $\chi^2$ | SIG. | LEVEL |
| --- | --- | --- | --- | --- | --- | --- |
|  | 53 (56.9) | 40 (43) | 93(100) | 2.55 | 0.11 | NS |
| AE Severity level | No. (%) | No. (%) | TOTAL | $\chi^2$ | SIG. | LEVEL |
| Low | 21 (22.5) | 7 (7.5) | 28 (30,1) | 7.122 | 0.28 | NS |
| Average | 17 (18.2) | 21 (22.5) | 38 (40,8) |  |  |  |
| High | 15 (16.1) | 12 (12.9) | 27 (29) |  |  |  |
| PSCI<br>(scale 24-104) | Value | Value | Value | T | SIG. | LEVEL |
|  | 52.67 | 52.96 | * | 0.12 | 0.91 | NS |
| PSPI<br>(scale 12-40) | Value | Value | Value | T | SIG. | LEVEL |
|  | 25.4 | 27.3 | * | 8.7 | 0.000 | S |
| S= Significant. NS= Not significant. * No value applies. |  |  |  |  |  |  |

Source: Chirinos, M. et al.

Table 3 shows the hypothesis test for the differences between public and private hospitals in relation to the study variables: AEP, Safety Culture Index, Safety Perception Index, and AE Severity.

According to Table 3, the PSCI was determined as unfavourable for both types of institution (PuH = 52.67 and PrH = 52.96) without any significant statistical differences (t = 0.12, sig = 0.91), since the culture perceived and expressed by health professionals is not linked to a particular type of hospital. The PSPI, however, displays a different behaviour according to the type of hospital (t = 8.7, sig = 0.000, PuH = 30.3, and PrH = 32.7), showing a moderately high perception in public hospitals, and high in private hospitals.

### III. Regression of variables: PSC and PSP, based on the level of severity and AEP identified

Figure 1 highlights the correlations identified between the variables. When we evaluate AEP and AE severity in relation to PSC (Figure 1), we see a strong and negative correlation (rho=-0.8), indicating that the behaviour of one variable is determined by the performance of the other (showing dependence). Therefore, the higher the prevalence of serious AEs, the lower the culture of the health professionals; similarly, if culture is favourable, AEP and AE severity are lower.

**Figure 1.**
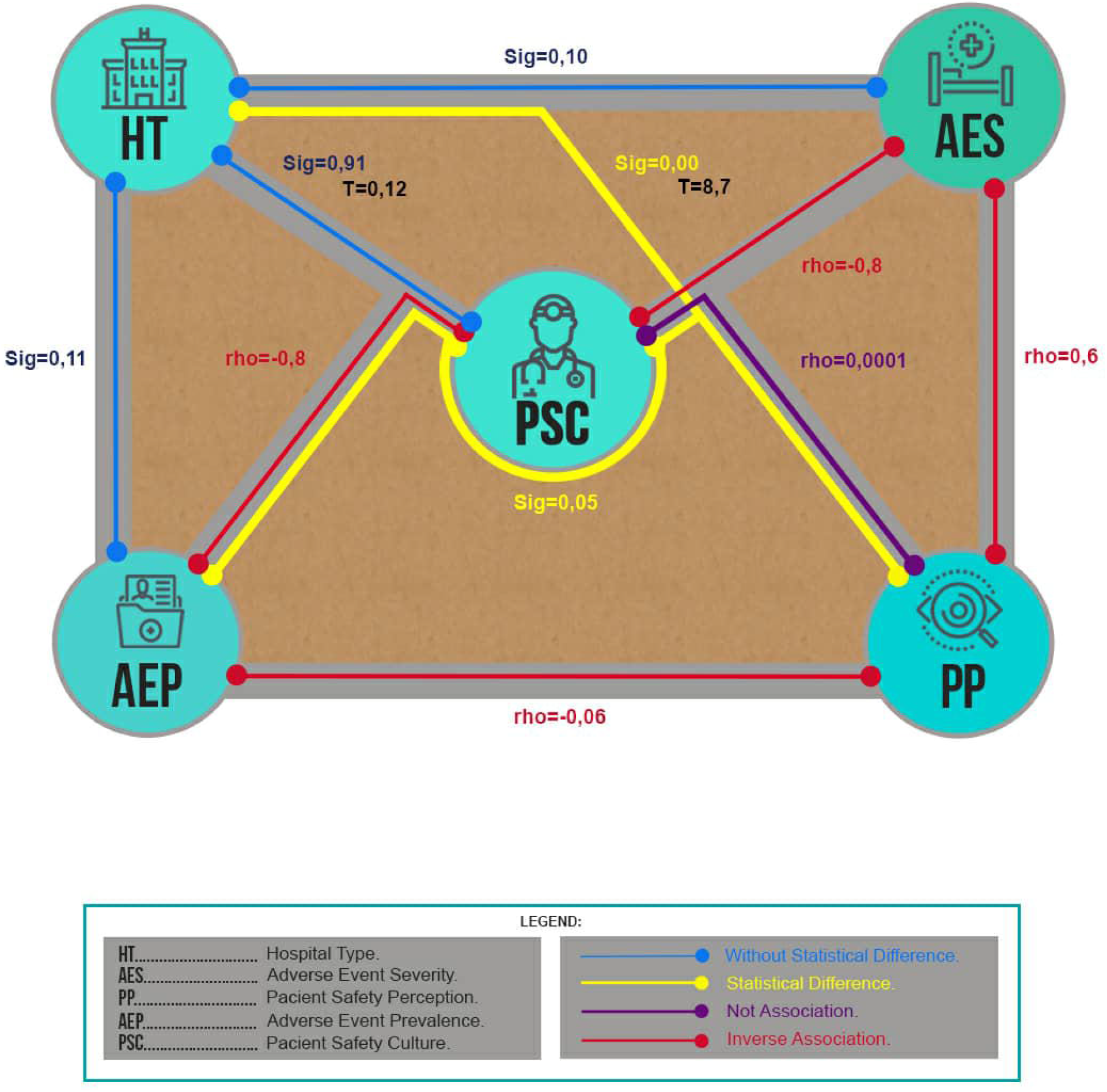
Proposed theoretical relational model showing the three aspects of patient safety and its results in our study.

An inverse relationship was found between the patient safety perceived by patients and AEP (rho = -0.06), that is, in the population studied, when there was a lower level of perception or observation, the occurrence of AEs was higher. In this study, the occurrence of AEs was high, while the perception of AEs was lower. On the other hand, severe AEs were observed the least, or the patients who experienced severe AEs actually detected them the least out of all the patients studied, and the most commonly observed AEs were mild (see Figure 1).

When analysing PSC and PSP, we can see that both variables are independent, with no correlation between them (rho = 0.0001). Contrary to the previous results, we have observed that the safety perceived by the patients was not related to the safety culture detected among the health professionals, as the patients did not detect the low level of culture that was identified among the professionals and it did not determine their outcomes. We therefore need to evaluate the other variables that allow us to define the patient’s perception (see Figure 1).

An inverse relationship was found in the sample studied between what the patients perceive about their safety and the prevalence of AEs: The occurrence of AEs was high, while the perception of AEs was low in both types of hospitals, with a moderately low PSCI (see Table 4). Furthermore, when relating these two variables, we also observed that, in general, in both types of hospital, patients had a moderately low perception of all types of AEs. However, it should be noted that even though severe AEs aggravated the patient’s condition in many cases, this kind was the least detected by the patients. Likewise, the AEs that were detected were mostly mild or moderate.

**Table 4.** Distribution of severity level of AE perceived and not perceived by patients, by hospital.

| Severity of AE | No. of AE not perceived by patients |  |  |  | Total AE not perceived |  | No. of AE perceived by patients. |  |  |  | Total AE perceived |  |
| --- | --- | --- | --- | --- | --- | --- | --- | --- | --- | --- | --- | --- |
|  | PuH |  | PrH |  |  |  | PuH |  | PrH |  |  |  |
|  | No. | % | No. | % | No. | % | No. | % | No. | % | No. | (%) |
| Low | 12 | 19.0 | 2 | 7.7 | 14 | 22.2 | 9 | 30.0 | 5 | 35.7 | 14 | 46.7 |
| Medium | 12 | 19.0 | 13 | 43.33 | 25 | 39.7 | 5 | 16.7 | 8 | 57.1 | 13 | 43.3 |
| High | 13 | 35.1 | 11 | 36.7 | 24 | 38.1 | 2 | 6.7 | 1 | 7.1 | 3 | 10.0 |
| Total | 37 | 69.8 | 26 | 65.0 | 63 | 100.0 | 16 | 30.2 | 14 | 46.7 | 30 | 100.0 |

### Distribution of the occurrence of Adverse Effects by severity and whether perceived by the patients or not

**Distribution of the occurrence of Adverse Effects by severity and whether perceived by the patients or not.**
| Severity of AE | No. of AE not perceived by patients. |  |  |  | Total AE not perceived. |  | No. of AE perceived by patients. |  |  |  | Total AE perceived by patients. |  |
| --- | --- | --- | --- | --- | --- | --- | --- | --- | --- | --- | --- | --- |
|  | PuH |  | PrH |  |  |  | PuH |  | PrH |  |  |  |
|  | No. | % | No. | % | No. | % | No. | % | No. | % | No. | (%) |
| Low | 12 | 19.0 | 2 | 7.7 | 14 | 22.2 | 9 | 30.0 | 5 | 35.7 | 14 | 46.7 |
| Medium | 12 | 19.0 | 13 | 43.33 | 25 | 39.7 | 5 | 16.7 | 8 | 57.1 | 13 | 43.3 |
| High | 13 | 35.1 | 11 | 36.7 | 24 | 38.1 | 2 | 6.7 | 1 | 7.1 | 3 | 10.0 |
| Total | 37 | 69.8 | 26 | 65.0 | 63 | 100.0 | 16 | 30.2 | 14 | 46.7 | 30 | 100.0 |

## DISCUSSION

Many researchers have individually studied the variables examined in this article: AEs,(1,3,6,8,25,26) PSC,(12,27) and PSP;(24) however, as far as we know, they have not integrated them into the same study. Focusing on what happens in the process of individual care and those involved in that care — patients, professionals, and institutions — is an innovative and unprecedented approach, which contributes methodological and theoretical value to the field of study. The study of how these variables are integrated arises from the theoretical and practical premise that patients(11) and health professionals, who interact in different ways in the care process, are in turn framed in other interconnected and interdependent institutional processes and protocols.(28,29)

The results of this study show that for patient safety to be more comprehensive and for the benefit to the patient to be effective and safe practices to be sustainable and internalized, improvement actions must focus on the care process and its actors, and address the variables in an integrated way.

Using the relational theoretical model, the main results of this study suggest that in the sample studied the nature of the relationship between the variables derives from the fact that the behaviour of one affects the other. Such is the case of the prevalence and severity of AEs (both types of hospital) (rho = 0.1), which present an inverse relationship with the culture of patient safety (rho = - 0.8) and in turn with the safety perceived by the patients (rho = -0.06). Hence, an increase in one leads to a decrease in the other in such a way that culture and perception are confirmed to be protective factors of patient safety and, consequently, can improve the rate of AEs as well as their severity and nature.

In this same sense, another relevant finding detected in this sample is the dissociation between what health professionals do and/or know in favour of patient safety (culture) and what patients perceive or observe regarding their safety (rho = 0.00). This finding suggests the need for further research that utilizes this supplementary information.

### Association between PSC perceived by health professionals and AEP, AE severity, and AE perception by patients

AEP linked to PSC is a field that has been widely researched;(29) however, the validity of some methodologies used to evaluate PSC has been questioned.(12,30,31) This was precisely one of the reasons why we decided to use a previously validated survey (HSOPSC, QPHS), which we consider to be complete, interesting, and useful, to propose a different qualitative form that creates indices for the analysis of each of the variables.

As expected in hospitals that present a low culture index, a high prevalence and severity of AEs was detected, maintaining an inverse relationship, as shown by some other studies,(12,30–32), and strengthening the theory that culture is an important criterion for maintaining a sustainable PSC.(9) Although it is stressed that there is insufficient evidence to show that safety culture directly impacts patient outcomes, there is a linear link between the two; these results add to the trend that there is a two-way relationship between the safety culture of health personnel and the clinical safety results of patients.(33)

On the other hand, in this sample, it is evident from the results of the patients who participated in their individual care processes that there is much ignorance about patient safety and its impact and role. An interesting study by Schwappah et al.(34) shows that 95% of patients and 78% of health workers agreed that both patient education and the willingness of patients to accept proposals from health professionals improve the patients’ perception of the care received (r = 1, 9, CI = 1.1 to 3.3, p = 0.034) and the desire of health professionals to improve their attitudes to patient safety. However, the study does not carry out a global assessment of the relationship between safety culture and perception.

### Association between patient perception of their clinical safety and the prevalence and severity of AEs

The inverse relationship found between AEP and patients’ perception of their safety revealed that levels of perception of AEs were low when AEs of any type of severity occurred; patient perception was lower when AEs were serious; and every kind of case generated insecurity, uncertainty and distrust regarding the care received.

With similar results to this research, authors such as Evans et al.(35) and Guijarro et al.(36) reported that regarding patients’ perception of their clinical safety, centres with a greater prevalence of AEs had a low PSPI. Although this finding may seem paradoxical, patient perception of this matter is affected by certain criteria or patient characteristics such as their level of observation, education and other aspects. Furthermore, as observed in this study and pointed out by other authors, the patient’s level of trust in the institution, the professionals and the care process itself varies depending on his/her perception of the latter. This reinforces the idea that a more active patient-user role needs to be promoted, as this would help to identify critical points in the care process and ensure that the appropriate measures are taken.(11) It is also a mitigating factor(37) and even a defence barrier(38) in the appearance of AEs and can contribute to institutional improvement, including AE reporting.(12,29,39). This idea should be explored more thoroughly in further studies.

With this assessment, as pointed out by Sammer et al.,(35) AEP is a strong predictor of the feeling of security. It is estimated that among the population studied at the public hospitals, 19 out of 100 patients admitted are likely to experience an AE, and seven of these would not detect having suffered any kind of AE. In private hospitals, 14 out of 100 patients admitted may suffer an AE, and five of these would not be aware of where the AE originated. Thus, in the population studied, when there was a lower level of perception or observation, the occurrence of AEs was higher.

The results also show that a significant percentage of severe AEs are poorly detected by patients (35.1% PuH, 36.7% PrH); This is attributed to the fact that they consider that everything that happens within a health institution is part of the pathological process that the patient goes through and that it is normal for these situations to happen, since patients do not have control, nor are they involved(40). Based on the experience in Australia, Evans et al. found that severity is an important predictor of how patients perceive safety,(35) that is, patients’ sense of security and trust in the hospital is defined by whether they or any of their relatives have experienced a serious AE when they were hospitalized (p <0.001; 95% CI).

The situations described respond, according to previous studies, to multiple failures in both institutional and individual healthcare processes,(41) a lack of communication with health professionals,(36) a culture of concealment of errors and/or characteristics specific to the population (e.g. educational level).(6,11) In this sense, there is an interesting study that concludes that there is likely to be a greater occurrence of AEs in patient populations with lower levels of education in safety concepts or a lack of awareness of the occurrence of unsafe acts in health institutions, which also act as a barrier to patients having an active participation.(6,36,41)

### Associations between the behaviour of public and private hospitals and the culture of health professionals, AEP and PSP

After analysing the prevalence, culture, and perception of AEs according to the type of hospital (public or private), no significant differences were observed between them, that is, the three variables behaved similarly in all of them. A systematic review revealed(16,19) that there is little evidence to show that private sector hospitals are more efficient or show greater clinical effectiveness than public hospitals in countries with medium and low levels of development. However, specific studies show the opposite, as in the case of Peru,(42) where the culture of safety is much higher in private than in public hospitals and Pakistan,(43,44) Nigeria and Malaysia,(45) where private sector hospitals enjoy greater user acceptance and satisfaction than public hospitals.

There were difficulties in finding any research similar to this study, in which the association between the culture of patient safety, the perception of patient safety, and the prevalence of AEs were evaluated in a single study, distinguishing between public and private hospitals. Therefore, and in order to minimize bias in national estimates, more in-depth research is needed in countries where prevalence, culture, and perception have already been studied.

## STRENGTHS AND LIMITATIONS

This study presents an association model of the variables of patient safety that represents the voice of the patients, the health professionals, and the institution where the processes took place, and therefore where the adverse events occurred, over a single time period. As a result, it examines the safety of the care process from the perspective of the patient, health professional and institution which participated in the process, thereby providing a new and integrated way of studying and analysing this topic as well as contributing to the overall knowledge of patient safety. With this aim in mind, three broad, specific studies were conducted prior to the present study to obtain data to underpin this investigative proposal and make it robust.

This study has some methodological limitations that should be addressed.

The selection of hospitals is not representative of hospitals in Venezuela generally. Due to resource constraints, we were only able to conduct the analysis in a sample of four high-level hospitals in one of the most populated regions of the country. Given the limited sample size, our findings cannot be generalized.

For the initial phases (1 and 2), a resistance to complete the instruments was evidenced, possibly because, as previously indicated, the care providers and patients have little awareness of this matter and/or there is a certain level of fear associated with it due to the threat of punitive measures. In order to avoid lower response rates in these cases, the professionals among whom resistance was detected were grouped together and given explanatory sessions about the instrument and its academic and investigative objective to ensure that they completed the questionnaire. This measure may have introduced differences when comparing the results with those of other similar studies.

## CONCLUSIONS

The main findings of this study provide a simultaneous assessment of the three main variables that affect patient safety: the culture of safety in the hospitals under study, AEP and the perception that patients have of safety. All of these variables were studied within the context of public or government sector institutions and the private sector. A relational theoretical model was used to establish the three aforementioned variables, with the addition of the severity with which AEs occur and the type of hospital, helping us to identify the dynamics of these variables.

This relational theoretical model is both systematic and flexible since it can be applied to other populations and can be used to determine a diagnosis of the organization or compare it with other organizations to allow objective and sustainable decision-making in this area.

The reflection on the behaviour of these variables in hospitals with different types of management (public and private) invites us to rethink the manner, purpose and scope of actions for implementing improvements in patient safety when patient-professional views and institutional processes are not integrated.

It is also vital to build a culture of quality patient safety in collaboration with the organizations, systems and processes that give patients a voice and which must be considered in conjunction with each other. Based on the results of this study, this consideration should be an active agent in the theoretical and practical models of error management.(38)

In order to validate the relationship model, it would be useful to apply it to other samples, as this would make it possible to observe different behaviours and provide an insight into how these variables interact in the institutions where it is applied.

## Data Availability

We declare that the data from which the results presented in this investigation are derived are available for reuse.
The available data refer to:
Adverse Events Prevalence, Patient safety Culture, Patient perception by patient safety.
Patient Safety Culture Index by hospital.
Patient Safety perception Index by hospital.
Adverse event severity by hospital.
Adverse event prevalence by hospital.
Adverse event characteristics.
This data is the result of another series of specific data derived from 3 instruments applied for the determination and study of each variable that corresponds to individually conducted studies, which are also available.
For the location of this data, the ORCID of the person responsible for managing this data is presented.
The reuse of this data is allowed under the conditions of other studies on the same database, which contribute to the body of knowledge of patient safety.

## Data availability statement

Data are available upon reasonable request and are not available publicly. Due to Venezuela’s Law on the Protection of Personal Data https://www.dlapiperdataprotection.com/index.html?t=law&c=VE, the data base used in the study remains under the safeguard of the authors and coauthors of the study. For permission to access the data, interested researchers are required to provide their name, institution, and purpose to avoid misuse of this sensitive data. Therefore, interested researchers may contact the head of data archiving (Cesar Montoya.)

## Ethics statement

This study was approved by the ethics committees at the Clinical Hospital of Maracaibo (Act N 07-15), the Rosario Hospital of Cabimas (Act N 06-16), Our Lady of Chiquinquirá Hospital of Maracaibo (Act N 0316), and the General Hospital of the South “Dr Pedro Iturbe” (Act N 25-06-16-1).

## Patient consent for publication

Not applicable.

## Acknowledgements

The authors would like to express their thanks to the Al Avedis Donabedian University Institute for its support and collaboration in carrying out this study, and to Louise Meadley for her support in the translation of this paper.

## Ethical considerations

- Validation of each institution by an ethics committee through the evaluation of research protocol. Hospital General del Sur “Dr. Pedro Iturbe”; Hospital Nuestra Señora de Chiquiquira; Hospital Clínico de Maracaibo; Hospital El Rosario, and College of Doctors of Zulia State.
- Application of informed consent from patients surveyed.

## Contributors

All authors made substantial contributions to the project conception, study design, data analysis and drafting of the paper. All authors have approved this version of the paper for publication.

## Copyright for authors

“The Corresponding Author has the right to grant on behalf of all authors and does grant on behalf of all authors, a worldwide licence to the Publishers and its licensees in perpetuity, in all forms, formats and media (whether known now or created in the future), to i) publish, reproduce, distribute, display and store the Contribution, ii) translate the Contribution into other languages, create adaptations, reprints, include within collections and create summaries, extracts and/or, abstracts of the Contribution, iii) create any other derivative work(s) based on the Contribution, iv) to exploit all subsidiary rights in the Contribution, v) the inclusion of electronic links from the Contribution to third party material where-ever it may be located; and, vi) license any third party to do any or all of the above.”

## Ethics approval

The studies that precede this investigation were approved by the ethics committees at the Clinic Hospital of Maracaibo (Act No. 07-15), The Rosario Hospital of Cabimas (Act No. 06-16), our Lady of Chiquinquirá Hospital of Maracaibo (Act No. 0316), General Hospital of the South “Dr. Pedro Iurbe” (Act No. 25-06-16-1).

